# Evaluation of anti-SARS-CoV-2 antibody testing in asymptomatic or mild COVID-19 patients in outbreak on a cruise ship

**DOI:** 10.1101/2021.03.10.21253064

**Authors:** Norihito Kaku, Fumitaka Nishimura, Yui Shigeishi, Rina Tachiki, Hironori Sakai, Daisuke Sasaki, Kenji Ota, Kei Sakamoto, Kosuke Kosai, Hiroo Hasegawa, Koichi Izumikawa, Koya Ariyoshi, Hiroshi Mukae, Jiro Yasuda, Kouichi Morita, Shigeru Kohno, Katsunori Yanagihara

## Abstract

**Background:** A few studies on antibody testing have focused on asymptomatic or mild coronavirus disease 2019 (COVID-19) patients with low initial anti-severe acute respiratory syndrome coronavirus 2 (SARS-CoV-2) antibody responses. Anti-SARS-CoV-2 antibody-testing performance was evaluated using blood samples from asymptomatic or mild COVID-19 patients.

**Methods:** Blood samples were collected from 143 COVID-19 patients during an outbreak on a cruise ship 3 weeks after diagnosis. Simultaneously, a second SARS-CoV-2 genetic test was performed. Samples stored before the COVID-19 pandemic were also used to evaluate the lateral flow immunochromatographic assay (LFA) and electrochemiluminescence immunoassay (ECLIA). Titers of anti-SARS-CoV-2 IgM and IgG antibodies against the nucleocapsid and spike proteins were measured using the enzyme-linked immunosorbent assay to compare false-negative- with positive-result samples.

**Results:** Sensitivity, specificity, positive-predictive, and negative-predictive values of LFA-detected IgM antibodies were 0.231, 1.000, 1.000, and 0.613, respectively; those of LFA-detected IgG antibodies were 0.483, 0.989, 0.972, and 0.601, respectively; and those of ECLIA-detected total antibodies were 0.783, 1.000, 1.000, and 0.848, respectively. IgM-, IgG-, and total-antibody positivity rates in the patients with negative results from the second genetic testing were 22.9%, 47.6%, and 72.4%, respectively. All antibody titers, especially those of the IgG antibody against nucleocapsid protein, were significantly lower in blood samples with false-negative results than in those with positive results.

**Conclusions:** These findings suggest that anti-SARS-CoV-2 antibody testing has lower performance in asymptomatic or mild COVID-19 patients than required in the guidelines, and situations in which it is useful are limited.

**Key points:** Anti-SARS-CoV-2 antibody testing in asymptomatic or mild COVID-19 patients is lower than the required clinical sensitivity, although it may be useful in patients at 3–4 weeks after symptom onset but with negative SARS-CoV-2 genetic test results.

## Introduction

The clinical indications for anti-severe acute respiratory syndrome coronavirus 2 (SARS-CoV-2) antibody testing are limited, although many methods have been developed. The detection of anti-SARS-CoV-2 IgG or total antibodies at 3–4 weeks after symptom onset may be useful in determining past infection in selected clinical situations; however, data in this context are limited. [3] On the other hand, anti-SARS-CoV-2 antibody testing may play an essential role in the public health response to coronavirus disease 2019 (COVID-19) and in understanding the outbreak dynamics of the COVID-19 pandemic.[4,5] However, a previous study reported that initial anti-SARS-CoV-2 antibody responses were lower in asymptomatic or mild COVID-19 patients than in severe COVID-19 patients.[1] Since the percentage of asymptomatic COVID-19 patients in large-sample studies was 43.0%–76.5%[2], it is necessary to elucidate the performance of anti-SARS-CoV-2 antibody testing in asymptomatic or mild COVID-19 patients.

We preserved blood samples of asymptomatic or mild COVID-19 patients collected from a large cruise ship that had experienced a COVID-19 outbreak. The cruise ship anchored at Nagasaki Port from the end of January 2020, and one of the 623 crew members complained of pneumonia symptoms and was diagnosed with COVID-19 in mid-April 2020. Thereafter, SARS-CoV-2 genetic testing was performed on all cruise ship crew members, and SARS-CoV-2 was detected in 144 individuals. However, most patients received care on the ship because they were asymptomatic or did not need inpatient treatment, such as oxygen administration. In this study, we evaluated the performance of anti-SARS-CoV-2 antibody testing using the lateral flow immunochromatographic assay (LFA) and electrochemiluminescence immunoassay (ECLIA) in blood samples from crew members and those collected from patients between November 2014 and August 2019. In addition, we measured the titers of several anti-SARS-CoV-2 antibodies and compared them between false-negative- and positive-result samples in anti-SARS-CoV-2 antibody testing using the enzyme-linked immunosorbent assay (ELISA).

## Methods

### Study design

In this study, asymptomatic or mild COVID-19 was defined as a cruise ship crew member who was not hospitalized when blood samples were collected. Blood samples were collected from 178 crew members in mid-May 2020, 3 weeks after the first SARS-CoV-2 genetic testing. Thirty-four blood samples were excluded from the final analysis due to negative results in the first SARS-CoV-2 genetic testing, and one blood sample was excluded because the patient was hospitalized at the time of sample collection (Fig. 1). Finally, 143 blood samples collected from crew members who had positive first SARS-CoV-2 genetic test results 3 weeks before blood samples were analyzed in this study. A total of 269 stored blood samples were collected from patients at Nagasaki University Hospital between November 2014 and August 2019. After deduplication, 174 blood samples from patients were used as negative controls in this study (Fig. 1). The blood samples were stored at − 80 °C until further antibody testing. Initially, anti-SARS-CoV-2 antibodies were detected using LFA and ECLIA by clinical technologists at Nagasaki University Hospital. Subsequently, to confirm which antibodies were influenced by LFA- and ECLIA-negative results in crew-member samples, IgM and IgG antibody titers against anti-SARS-CoV-2 spike-protein (SP) and nucleocapsid protein (NP) were measured using ELISA at Nagasaki University Hospital by Cellspect Co., Ltd, researchers. Anti-SARS-CoV-2 antibodies were detected without notification of each result. Results from the second SARS-CoV-2 genetic testing using real-time reverse transcription polymerase chain reaction RT-PCR[6] testing were obtained from the database. The second genetic testing was performed on nasopharyngeal samples collected from cruise ship crew members at approximately the same time as blood samples were collected.

**Figure 1.**
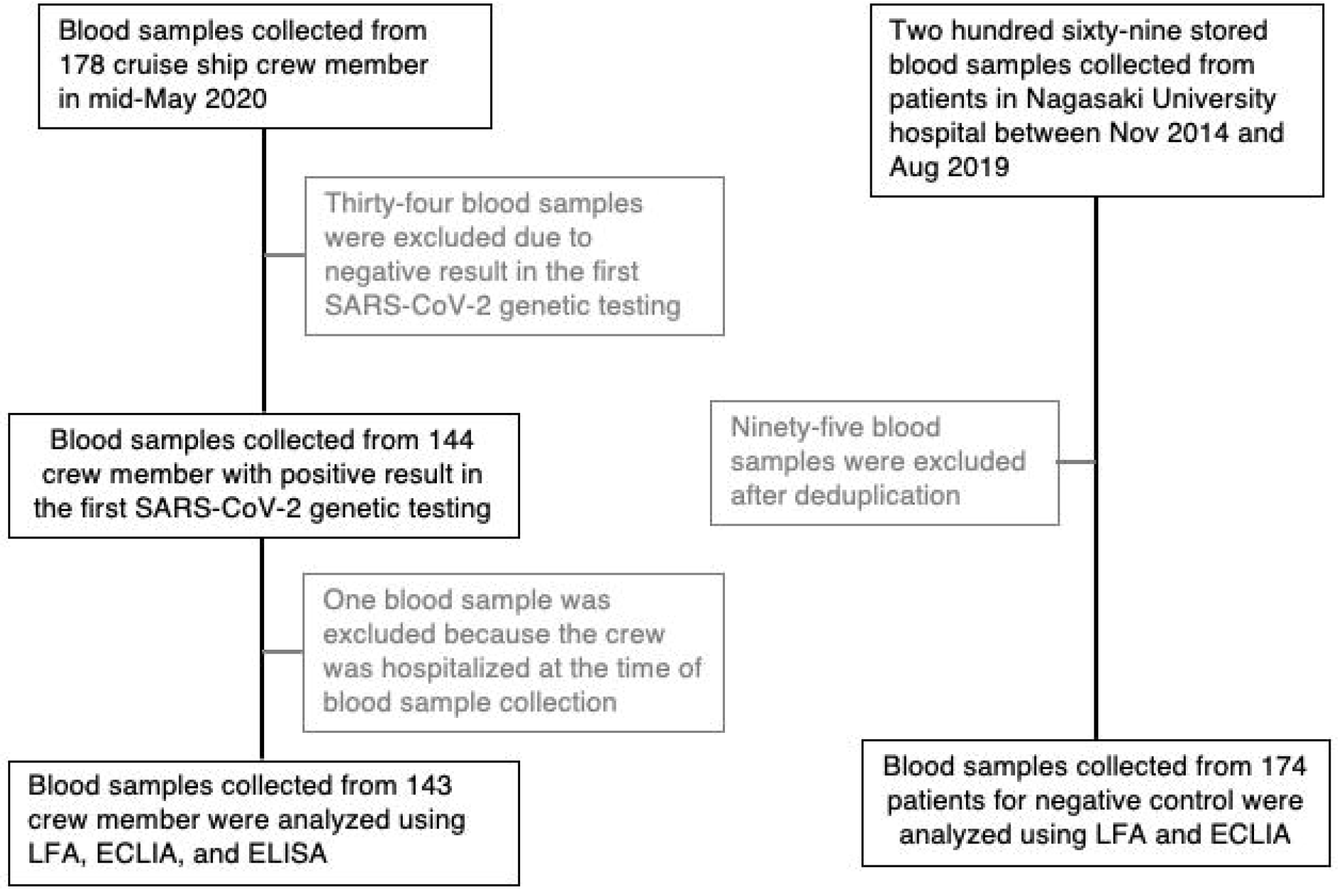
Participant flow diagram. SARS-CoV-2, severe acute respiratory syndrome coronavirus 2; LFA, lateral flow immunochromatographic assay; ECLIA, electrochemiluminescence immunoassay; ELISA, enzyme-linked immunosorbent assay

### Detection of anti-SARS-CoV-2 antibodies

SARS-CoV-2 IgM and IgG were detected using the SARS-CoV-2 Antibody IgM/IgG LFA testing kit (RF-NC001 and RF-NC002, Kurabo Industries, Ltd., Osaka, Japan) according to the manufacturer’s instructions. The results from tests using these kits were assessed by two clinical technologists. Anti-SARS-CoV-2 total antibodies were detected by Elecsys Anti-SARS-CoV-2 (Roche Diagnostics K.K., Tokyo, Japan) using a Cobas e801 analytical unit for immunoassay tests (Roche Diagnostics K.K., Tokyo, Japan) according to the manufacturer’s instructions. The judgment criteria for ECLIA were as follows: positive, cutoff index (COI) ≥ 1.0; negative, COI < 1.0. To measure anti-SARS-CoV-2 IgM and IgG antibodies against SP (QuaResearch COVID-19 human IgM/IgG ELISA kit, RCOEL961N, Cellspect Co., Ltd., Iwate, Japan) and those against NP (QuaResearch COVID-19 human IgM/IgG ELISA kit, RCOEL961, Cellspect Co., Ltd., Iwate, Japan), ELISA was performed by Cellspect Co., Ltd. The antigen proteins immobilized in the ELISA kit were recombinant SP (16-1213 AA) of SARS-CoV-2 expressed in mammalian cells and recombinant NP (1-419 AA) of SARS-CoV-2 expressed in *Escherichia coli*. To measure IgM and IgG antibodies against SP, serum or plasma samples were diluted 1:1000 in 1% bovine serum albumin/phosphate-buffered saline with Tween detergent (PBST); to measure IgM and IgG antibodies against NP, serum or plasma samples were diluted 1:1000 in 2% non-fat milk/PBST. The absorbance was read at 450 nm using an automatic ELISA system (QRC5LB925, Cellspect Co., Ltd., Iwate, Japan) according to the manufacturer’s instructions. The cutoff value determined in a previous study was used to interpret the ELISA results. [7] The cutoff values for anti-SARS-CoV-2 IgM and IgG antibody titers against SP were set at 0.25 and 0.26 and those against NP at 0.4 and 0.7, respectively. [7]

### Statistical analysis

All statistical analyses were performed using EZR version 1.53 (Saitama Medical Center, Jichi Medical University, Saitama, Japan), which is a graphical user interface for R (version 4.0.3; R Foundation for Statistical Computing, Vienna, Austria). Fisher’s exact test or the chi-square test was used to compare categorical variables and the Mann–Whitney U test to compare continuous variables. The statistical significance level was set at P <0.05. The sensitivity (Se), specificity (Sp), positive predictive value (PPV), and negative predictive value (NPV) of the antibody testing were calculated with 95% confidence intervals (95% CI). Spearman’s rank-order correlation was used to evaluate the correlation between anti-SARS-CoV-2 antibody titers in ELISA and COIs in ECLIA.

### Ethics

This retrospective study was approved by the Ethics Committee of Nagasaki University Hospital (20052101) and was registered in the UMIN Clinical Trials Registry (UMIN000040402). Data regarding SARS-CoV-2 RT-PCR results and blood samples were anonymized and individually numbered when they were collected from the cruise ship. Blood samples for the negative controls were stored anonymously at the Department of Laboratory Medicine, Nagasaki University Hospital.

### Data availability

Raw data were generated at Nagasaki University Hospital. Derived data supporting the findings of this study are available from the corresponding author upon request.

## Results

### Performance of LFAand ECLIA in detecting anti-SARS-CoV-2 antibodies

Anti-SARS-CoV-2 IgM and IgG antibodies were detected using LFA in 33 (23.1%) and 69 (48.3%) blood samples collected from crew members, respectively (Figs. 2A and 2B). The blood-sample positivity rate for LFA-detected anti-SARS-CoV-2 IgG antibodies was significantly higher than that for IgM (p < 0.001). Anti-SARS-CoV-2 IgG antibodies were detected in all blood samples, with positive results for LFA-detected IgM antibodies. There was no positive result for LFA-detected anti-SARS-CoV-2 IgM antibodies in the negative controls, whereas there were two positive results for anti-SARS-CoV-2 IgG antibodies in the negative controls. The Se, Sp, PPV, and NPV of LFA-detected anti-SARS-CoV-2 IgM antibodies were 0.231, 1.000, 1.000, and 0.613, respectively (Fig. 2A). The Se, Sp, PPV, and NPV of LFA-detected anti-SARS-CoV-2 IgG antibodies were 0.483, 0.989, 0.972, and 0.601, respectively (Fig. 2B).

**Figure 2.**
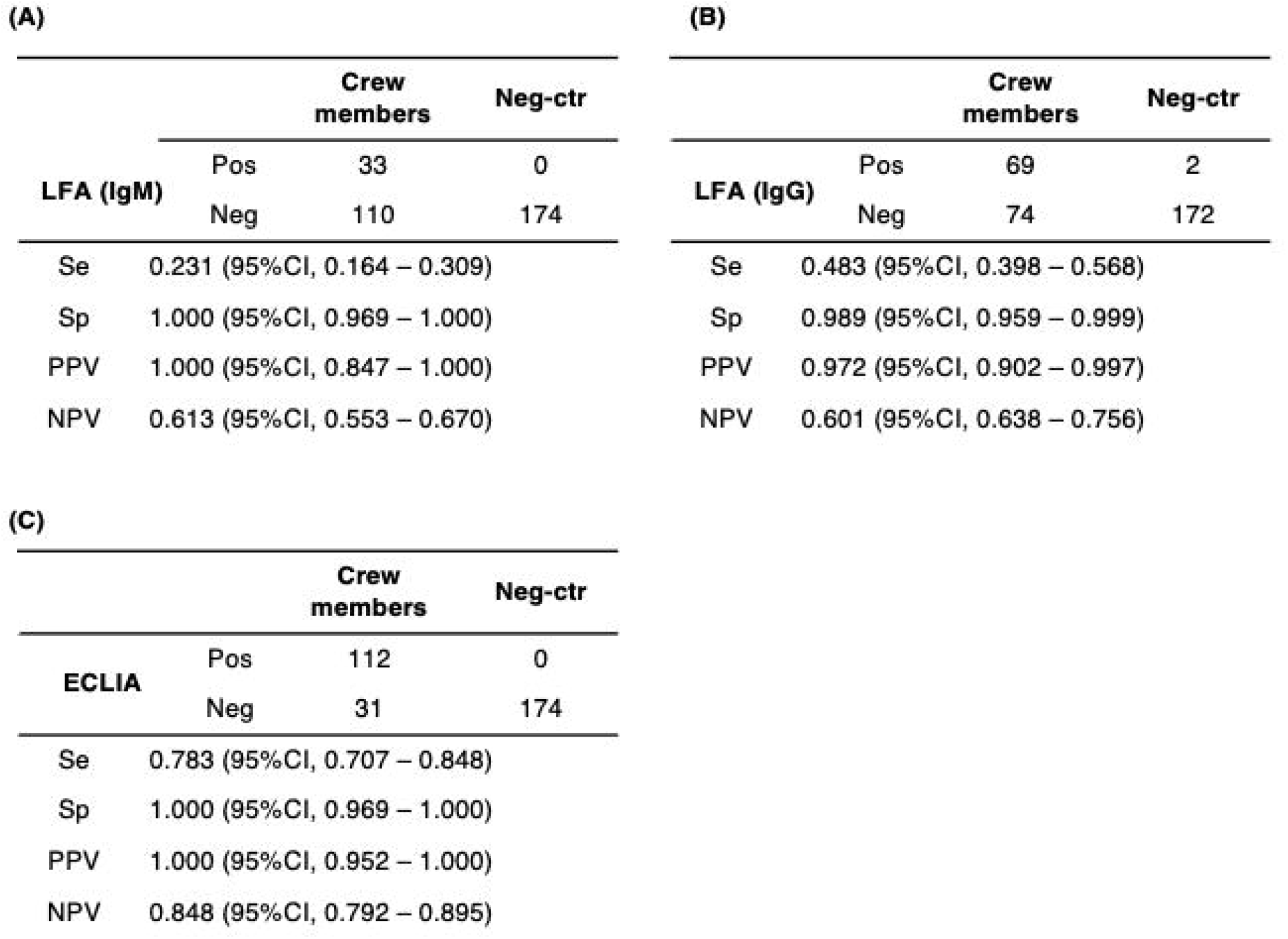
Performance of LFA and ECLIA in detecting anti-SARS-CoV-2 antibodies. Anti-SARS-CoV-2 IgM (A) and IgG (B) antibodies were detected using a lateral flow immunochromatographic assay (LFA), and anti-SARS-CoV-2 total antibodies were detected using an electrochemiluminescence immunoassay (ECLIA). Blood samples were collected from cruise ship crew members who had positive results from the first SARS-CoV-2 genetic testing 3–4 weeks before blood collection. Neg-Ctr, blood samples for negative controls that were collected from patients at Nagasaki University Hospital between November 2014 and August 2019; Se, sensitivity; Sp, specificity; PPV, positive predictive value; NPV, negative predictive value

Anti-SARS-CoV-2 total antibodies were detected using ECLIA in 112 (78.3%) blood samples collected from the crew (Fig. 2C). There was no positive result for ECLIA-detected anti-SARS-CoV-2 total antibodies in the negative controls. Anti-SARS-CoV-2 total antibodies were detected using ECLIA in all blood samples, with positive IgG antibody results detected by LFA. The blood-sample positivity rate for ECLIA-detected anti-SARS-CoV-2 total antibodies was significantly higher than that for LFA-detected IgM and IgG antibodies (p < 0.001). The Se, Sp, PPV, and NPV of ECLIA-detected anti-SARS-CoV-2 total antibodies were 0.783, 1.000, 1.000, and 0.848, respectively (Fig. 2C).

Among 143 COVID-19 patients, 105 (73.4%) tested negative in the second SARS-CoV-2 genetic testing using nasopharyngeal samples collected at approximately the same time as blood samples were collected. The positive rates of LFA-detected anti-SARS-CoV-2 IgM and IgG antibodies and ECLIA-detected total antibodies in the crew with positive results from the second SARS-CoV-2 genetic testing were 23.7% (9/38), 50.0% (19/38), and 94.7% (36/38), respectively. The positivity rates in crew members with negative results from the second SARS-CoV-2 genetic testing were 22.9% (24/105), 47.6% (50/105), and 72.4% (76/105), respectively. The positivity rate for ECLIA-detected anti-SARS-CoV-2 total antibodies was significantly higher in crew members with positive results from the second SARS-CoV-2 genetic testing than that in crew members with negative results (p = 0.003).

### Anti-SARS-CoV-2 antibody titers measured using ELISA in COVID-19 patients

Anti-SARS-CoV-2 SP IgM and IgG and anti-SARS-CoV-2 NP IgM and IgG antibody titers in blood samples collected from crew members were measured using ELISA. The median anti-SARS-CoV-2 IgM and IgG antibody titers against SP were 0.10 (0.07–0.47) and 0.27 (0.07–0.71), and those against NP were 0.18 (0.10–1.08) and 0.56 (0.18–2.82), respectively. The positivity rate of anti-SARS-CoV-2 IgM and IgG antibodies against SP in COVID-19 patients was 1.4% (2/143) and 31.5% (45/143), and those against NP were 3.5% (5/143) and 44.8% (64/143), respectively.

### Comparison of anti-SARS-CoV-2 antibody titers between samples with false-negative and those with positive LFA results

Antibody titers were compared between blood samples with false-negative results and those with positive LFA results (Fig. 3). Antibody titers for all antibodies were significantly higher in blood samples with positive results than in those with false-negative results (Fig. 3A-D). COI in ECLIA was also significantly higher in blood samples with positive results than in those with false-negative results (Fig. 3E). The positivity rates for ELISA-detected anti-SARS-CoV-2 IgM and IgG antibodies against SP in samples with false-negative results were 0% (0/74) and 16.2% (12/74), and those against NP were 1.4% (1/74) and 9.5% (7/74), respectively. The positivity rates for ELISA-detected anti-SARS-CoV-2 IgM and IgG antibodies against SP in samples with positive results were 2.9% (2/69) and 47.8% (33/69), and those against NP were 5.8% (4/69) and 82.6% (57/69), respectively. The positivity rates for anti-SARS-CoV-2 IgG antibodies against SP and NP were significantly lower in samples with false-negative results than in those with positive results (both p<0.001).

**Figure 3.**
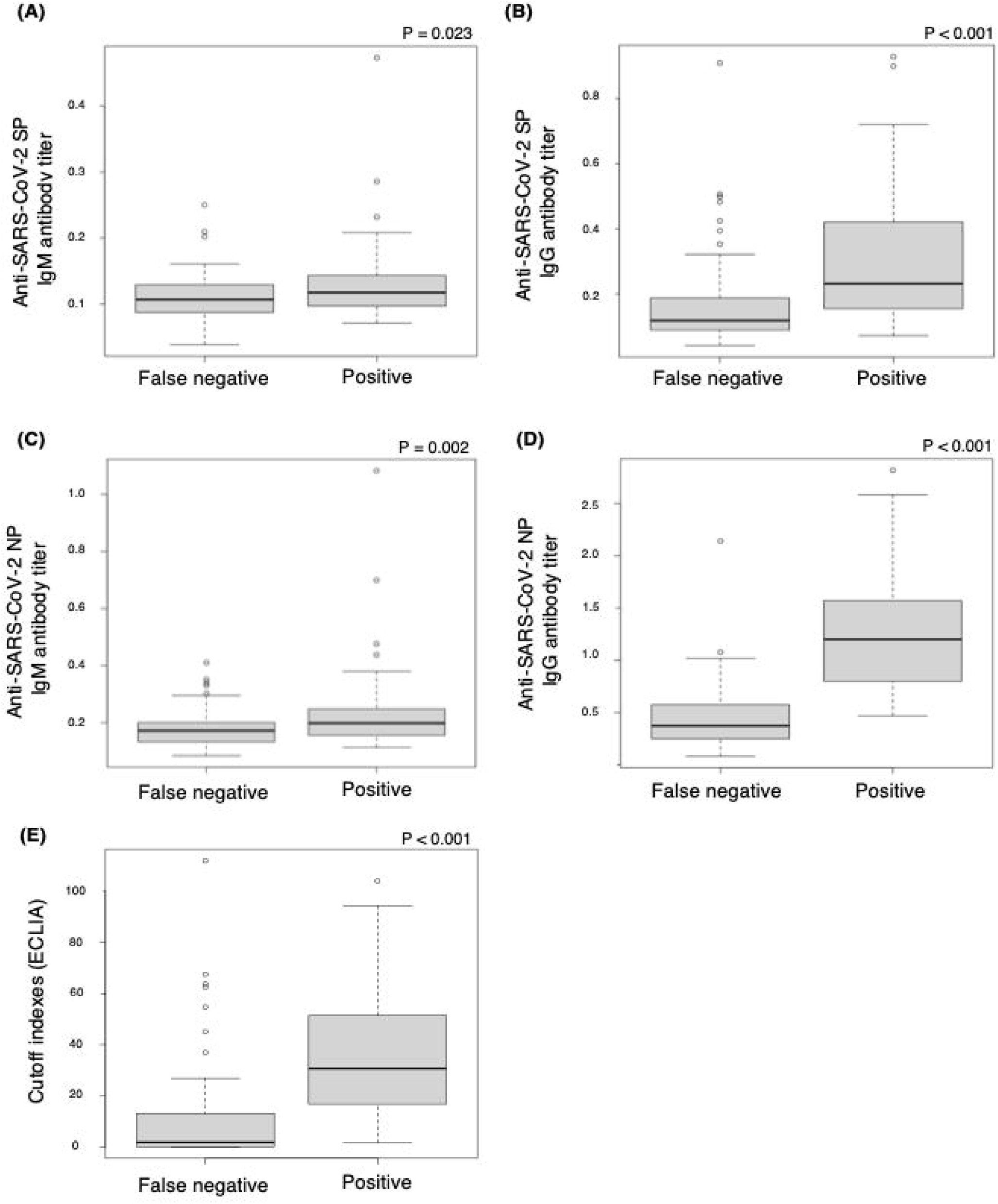
Comparison of anti-SARS-CoV-2 antibody titers between samples with false-negative and those with positive LFA results. Anti-SARS-CoV-2 SP IgM (A) and IgG (B) and anti-SARS-CoV-2 NP IgM (C) and IgG (D) antibody titers in blood samples collected from 143 cruise ship crew members who had positive first SARS-CoV-2 genetic test results were measured using ELISA. Antibody titers were significantly lower in blood samples with false-negative LFA results (n=74) than in those with positive LFA results (n=69). The cutoff index in ECLIA was also significantly lower in blood samples with false-negative LFA results than in those with positive LFA results. SP, spike protein; NP, nucleocapsid protein; ELISA, enzyme-linked immunosorbent assay; LFA, lateral flow immunochromatographic assay; ECLIA, electrochemiluminescence immunoassay.

### Comparison of anti-SARS-CoV-2 antibody titers between samples with false-negative and those with positive ECLIA results

Antibody titers were compared between blood samples with false-negative results and those with positive ECLIA results (Fig. 4). Antibody titers for all antibodies were significantly higher in blood samples with positive results than in those with false-negative results (Figs. 4A–D). The positivity rates for ELISA-detected anti-SARS-CoV-2 IgM and IgG antibodies against SP in samples with false-negative results were 0% (0/31) and 9.7% (3/31), and those against NP were 3.2% (1/31) and 0% (0/31), respectively. The positivity rates for ELISA-detected anti-SARS-CoV-2 IgM and IgG antibodies against SP in samples with positive results were 1.8% (2/112) and 37.5% (42/112), and those against NP were 3.6% (4/112) and 57.1% (64/112), respectively. The positivity rates for anti-SARS-CoV-2 IgG antibodies against SP and NP were significantly lower in samples with false-negative results than in those with positive results (p=0.004 and p<0.001, respectively).

**Figure 4.**
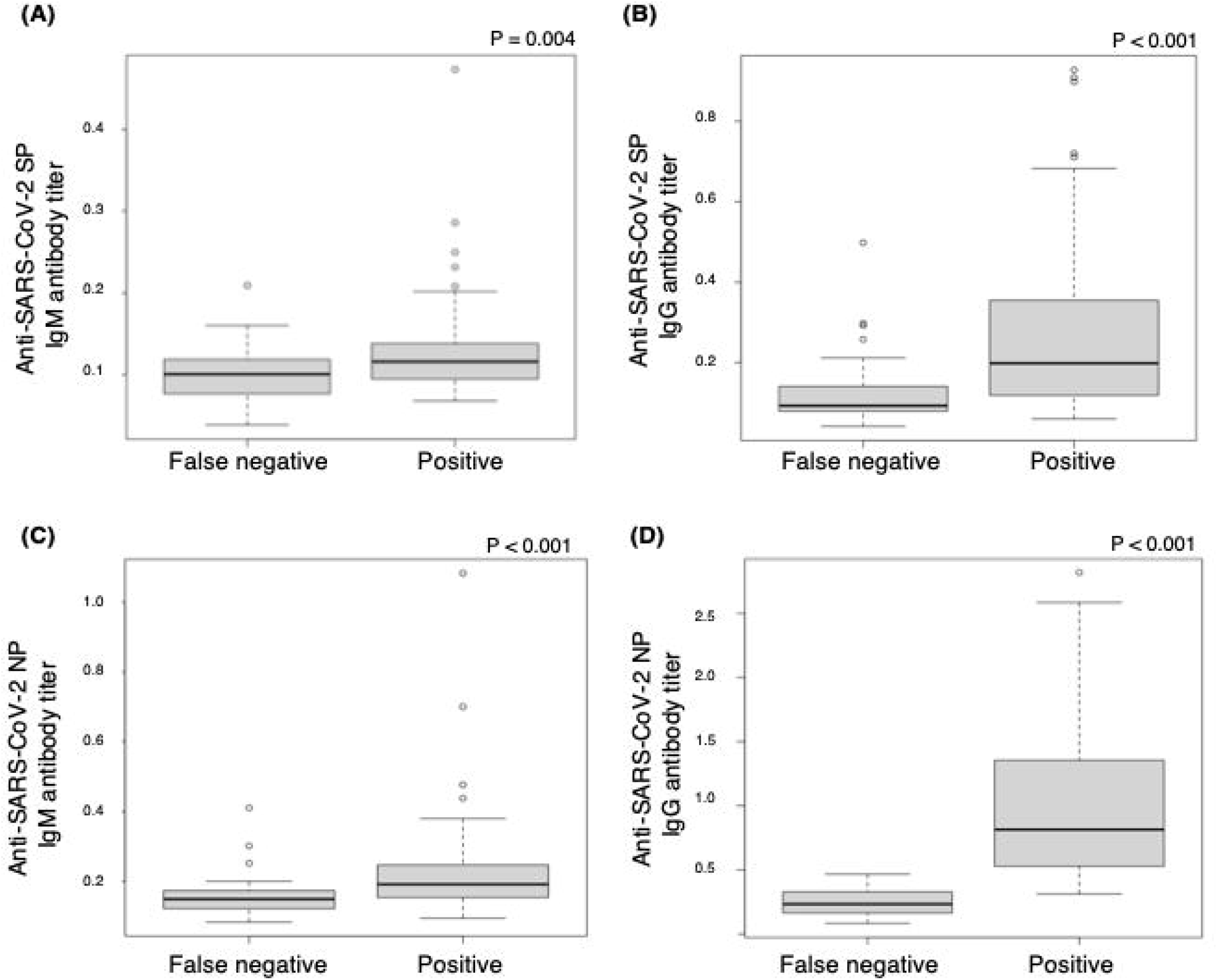
Comparison of anti-SARS-CoV-2 antibody titers between samples with false-negative and those with positive ECLIA results. SARS-CoV-2 SP IgM (A) and IgG (B) and SARS-CoV-2 NP IgM (C) and IgG (D) antibody titers in blood samples collected from 143 cruise ship crew members who had positive first SARS-CoV-2 genetic test results were measured using ELISA. Antibody titers were significantly lower in blood samples with false-negative ECLIA results (n=31) than in those with positive ECLIA results (n=112). SP, spike protein; NP, nucleocapsid protein; ELISA, enzyme-linked immunosorbent assay; ECLIA, electrochemiluminescence immunoassay.

The correlation between anti-SARS-CoV-2 antibody titers in ELISA and COI in ECLIA among blood samples collected from crew members was also evaluated (Fig. 5). The correlation coefficients between COI in ECLIA and anti-SARS-CoV-2 IgM and IgG antibodies against SP were 0.324 and 0.398, and those against NP were 0.332 and 0.812, respectively.

**Figure 5.**
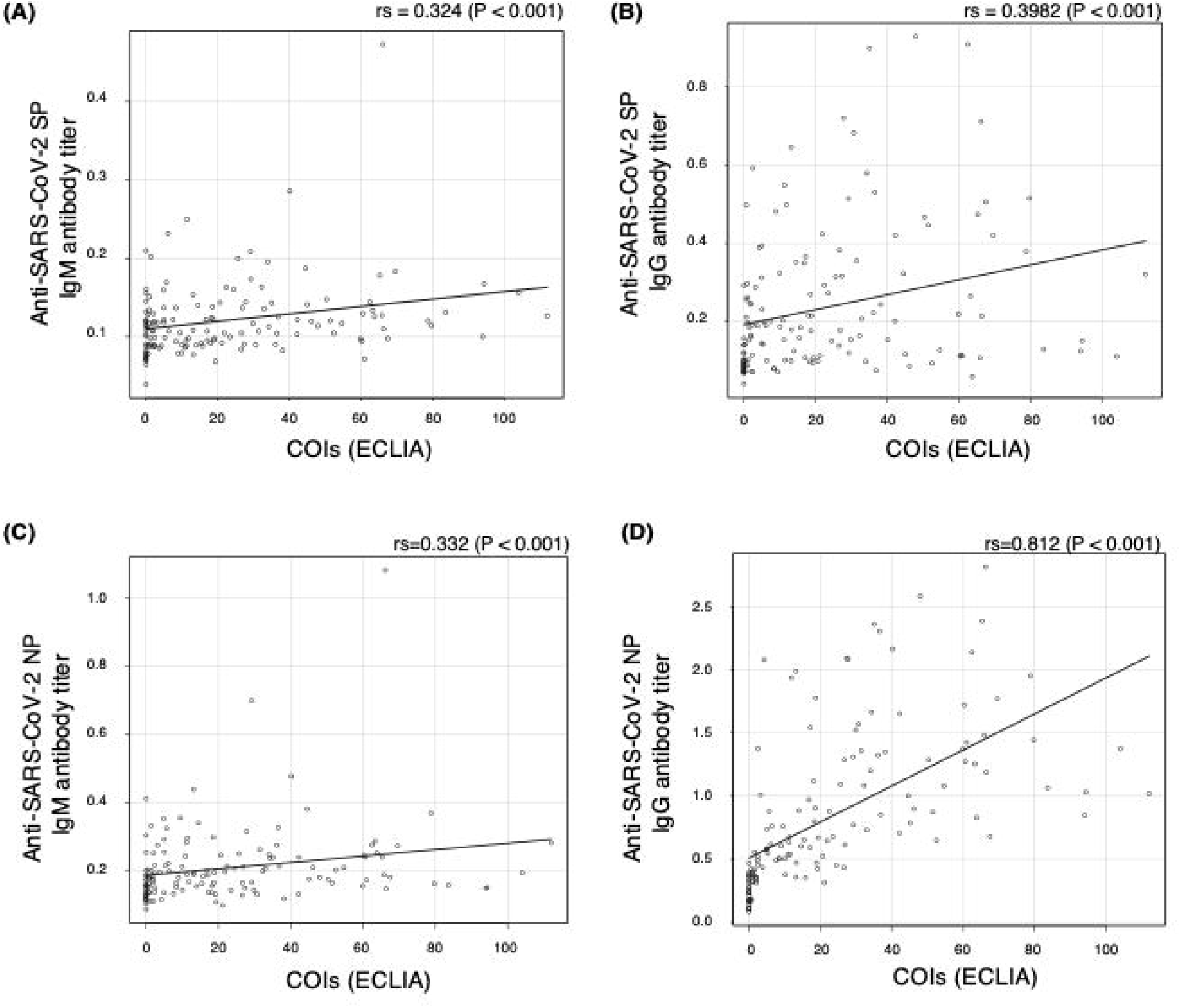
Correlation between anti-SARS-CoV-2 antibody titers in ELISA and COI in ECLIA. The correlation between SARS-CoV-2 antibody titers in ELISA and cutoff indexes (COIs) in ECLIA in blood samples collected from 143 cruise ship crew members who had positive first SARS-CoV-2 genetic test results were evaluated using Spearman’s rank-order correlation. SP, spike protein; NP, nucleocapsid protein; ELISA, enzyme-linked immunosorbent assay; ECLIA, electrochemiluminescence immunoassay.

## Discussion

This study revealed the performance of LFA and ECLIA in detecting anti-SARS-CoV-2 antibodies in asymptomatic or mild COVID-19 patients. The sensitivities of LFA for both anti-SARS-CoV-2 IgM and IgG antibodies were low (0.231 and 0.483, respectively) in this study. LFA’s sensitivity in this study was lower than that reported in two previous studies. One previous study reported IgM- and IgG-antibody positivity in all 24 COVID-19 patients.[8] In another study using blood samples collected from 12 COVID-19 patients, including two asymptomatic (16.7%) and seven mild patients (58.3%), the kit’s sensitivity for IgM and IgG antibodies was 0.750 and 0.727, respectively. [9] ECLIA’s sensitivity for anti-SARS-CoV-2 total antibodies was 0.783 in this study, which was higher than that of LFA. However, ECLIA’s sensitivity in this study was lower than that reported in previous studies using the same ECLIA kit; the sensitivity in previous studies was 0.920–0.995 [10–12]. One study reported that ECLIA’s sensitivity in all patients was 0.920–0.927; however, ECLIA’s sensitivity in 6 (4.0%) asymptomatic patients and 37 (24.8%) mild patients was reported to be approximately 0.800.[10] The results of this study demonstrated that ECLIA’s sensitivity in detecting anti-SARS-CoV-2 antibodies was relatively low in asymptomatic or mild COVID-19 patients. Considering the results of this study, the sensitivities of LFA and ECLIA in asymptomatic or mild COVID-19 patients were lower than the required clinical sensitivity in the guidelines. [3]

In contrast to sensitivity, the specificities of LFA and ECLIA were very high in this study. These results were consistent with those of previous studies using the same kits [10–14]. Although there were no samples with false-positive results for LFA-detected anti-SARS-CoV-2 IgM antibodies and ECLIA-detected total antibodies, there were two samples with false-positive results for LFA-detected SARS-CoV-2 IgG antibodies. A previous study using the same LFA kit reported SARS-CoV-2 IgG antibody positivity in 57% (4/7) of patients with human common cold coronavirus pneumonia.[8] The results from the previous study indicated that the LFA kit used in this study had cross-reactivity with antibodies against human common cold coronavirus. Although the false-positivity rate was very low in this study, the LFA kit may not be suitable for screening COVID-19 patients or for COVID-19 serological surveillance in a population with a low prevalence of COVID-19 because specificity is especially important in such situations.[3,15] In this study, 73.4% of COVID-19 patients tested negative in the second SARS-CoV-2 genetic testing using nasopharyngeal samples collected at approximately the same time as blood samples were collected. However, LFA and ECLIA detected anti-SARS-CoV-2 antibodies in 47.6% and 72.4% of patients with negative results in the second SARS-CoV-2 genetic testing, respectively. These data support the recommendation in the guidelines, [3] although it is necessary to consider the possibility of false-negatives in anti-SARS-CoV-2 antibody testing.

Anti-SARS-CoV-2 IgM and IgG antibody titers against SP and NP were measured using ELISA in this study. All antibody titers were significantly lower in samples with false-negative results than in those with positive results in both LFA and ECLIA. Among the antibodies, anti-SARS-CoV-2 IgG antibodies against SP and NP were noticeably divided between blood samples with positive and those with false-negative results in both methods. Additionally, the anti-SARS-CoV-2 IgG antibody titer against NP in ELISA was strongly correlated with COI in ECLIA. The ECLIA kit used in this study detects total antibodies (IgG, IgA, and IgM) against NP [16,17]; however, our data indicated that a low titer of anti-SARS-CoV-2 antibody against NP contributed to the false-negative results in asymptomatic and mild COVID-19 patients. A previous study reported that a combination of anti-SARS-CoV-2 antibodies against NP and SP increased the percentage of positive results.[18] In fact, the anti-SARS-CoV-2 antibody titer against SP in ELISA had a weaker correlation with COI in ECLIA in this study. However, anti-SARS-CoV-2 IgM and IgG positivity rates were also very low in COVID-19 patients with false-negative ECLIA results. Since all anti-SARS-CoV-2 antibody testing that was granted emergency use authorization by the United States Food and Drug Administration targeted SP and/or NP, it is currently difficult to improve the sensitivity of anti-SARS-CoV-2 antibody testing in asymptomatic or mild COVID-19 patients.

This study has several limitations. First, although all COVID-19 patients analyzed in this study were either asymptomatic or mild, we did not know their distribution. Because the positivity rates of ELISA-detected anti-SARS-CoV-2 IgG antibodies against SP and NP were lower than those in mild COVID-19 patients in a previous study[7], it is expected that the percentage of asymptomatic COVID-19 patients was higher than that of mild COVID-19 patients. Second, only one kit each of LFA, ECLIA, and ELISA was used in this study. Thus, other kits could have possibly had higher sensitivity in asymptomatic and mild COVID-19 patients than those used in this study. However, since the ECLIA used in this study demonstrated good sensitivity compared to other ECLIA kits,[10–12] it may be difficult to improve sensitivity by using others. Third, the time-course of the anti-SARS-CoV2 antibody testing was not evaluated in this study. This study focused on the time point that was reported as one of the best times for anti-SARS-CoV2 antibody testing [7,9,10]; however, it is possible that the optimal time for anti-SARS-CoV-2 antibody testing in asymptomatic or mild COVID-19 patients varies. Finally, the neutralizing antibodies were not evaluated. A previous study reported that the results of the ECLIA kit used in this study were correlated with SARS-CoV-2 neutralization; nevertheless, it also reported that the kit had poor negative-percent agreement for SARS-CoV-2 neutralization. Therefore, further investigation is required to confirm whether COVID-19 patients with false-negative results have neutralizing antibodies.

In conclusion, the findings of this study corroborate the recommendations of the guidelines. [3] They suggest that the sensitivity of anti-SARS-CoV-2 antibody testing in asymptomatic or mild COVID-19 patients is lower than the required clinical sensitivity. In addition, it may be difficult to improve anti-SARS-CoV-2 antibody testing sensitivity at present because anti-SARS-CoV-2 IgM and IgG antibody titers against SP and NP were very low in COVID-19 patients with false-negative results. On the other hand, anti-SARS-CoV-2 antibody testing may be useful in limited contexts, such as in patients at 3–4 weeks after symptom onset but with negative SARS-CoV-2 genetic test results.

## Acknowledgments

We would like to thank Editage (www.editage.com) for English language editing

